# Improving Immediate Postpartum Detection of Obstetric Anal Sphincter Injury: Added Diagnostic Value of Impedance Spectroscopy Following Clinical Assessment

**DOI:** 10.64898/2026.09.09.26362618

**Authors:** Małgorzata Uchman-Musielak, Katarzyna Kleczkowska, Iwona Sudoł-Szopińska, Katarzyna Borycka

## Abstract

**Objective:** To evaluate the added diagnostic value of immediate postpartum impedance spectroscopy (IS) when used alongside clinical assessment for obstetric anal sphincter injury (OASI), using subsequent three-dimensional endoanal ultrasound (3D EAUS) as the anatomical reference assessment.

**Methods:** This prospective observational cohort study included women following vaginal birth with at least one predefined OASI risk factor or clinical uncertainty regarding sphincter injury. Participants underwent postpartum clinical assessment, including digital rectal examination and senior obstetric reassessment when required, followed by immediate postpartum IS. Women who subsequently completed 3D EAUS were included in the paired diagnostic analysis. Diagnostic performance, agreement with EAUS and clinically relevant discordant findings were evaluated.

**Results:** Of 151 women enrolled, 70 completed EAUS and were included in the paired analysis; 29/70 (41.4%) were OASI-positive on the EAUS-based reference assessment. IS had a sensitivity of 86.2% (95% CI 69.4–94.5), specificity of 92.7% (95% CI 80.6–97.5) and overall accuracy of 90.0% (95% CI 80.8–95.1), compared with 75.9% (95% CI 57.9–87.8), 70.7% (95% CI 55.5–82.4) and 72.9% (95% CI 61.5–81.9), respectively, for clinical assessment. In paired analysis, IS was correct and clinical assessment incorrect in 17 women, whereas clinical assessment was correct and IS incorrect in five (McNemar P=0.017). Seven women with confirmed OASI were not identified clinically, and all seven generated an IS REFER result. Conversely, among 12 women clinically diagnosed with OASI who underwent primary repair but were subsequently classified as OASI-negative, 10 (83.3%) generated an IS PASS result. IS was performed within the first hour postpartum in 94.3% of women, and 70.0% of examinations were performed by midwives.

**Conclusions:** Immediate postpartum IS provided additional objective information complementary to clinical assessment, particularly when clinical findings did not reliably identify or exclude sphincter injury. These findings support prospective evaluation of IS within postpartum diagnostic pathways to determine whether its use reduces diagnostic errors and improves clinically meaningful outcomes.

## Introduction

Obstetric anal sphincter injury (OASI) is an important cause of anal incontinence following vaginal birth. Recognition at delivery is essential to allow appropriate primary repair and subsequent follow-up. However, OASI may be missed or incorrectly classified during the initial postpartum examination.^1–3^

International guidance recommends systematic examination after vaginal birth, including digital rectal examination (DRE), to identify OASI.^4^ Nevertheless, clinical assessment remains dependent on the examiner’s ability to recognize disrupted sphincter anatomy in the acute postpartum setting.

Ultrasound studies confirm that clinical and anatomical findings do not always agree. In a recent meta-analysis, 20% of women in studies reporting both clinical and ultrasound findings had anal sphincter trauma detected on ultrasound that had not been reported clinically at childbirth.^5^ Internal anal sphincter injury may be particularly under-recognized during clinical assessment.^6^

Endoanal ultrasound (EAUS) provides detailed anatomical assessment of the external and internal anal sphincters and is the established reference imaging modality for structural assessment of the anal sphincter complex.^7^ Its routine use immediately after birth, however, is difficult. Immediate postpartum studies have reported challenges in image interpretation, while access to appropriate equipment and experienced operators may be limited.^8^ This leaves a diagnostic gap between routine bedside examination, when decisions about further assessment and treatment are made, and specialist anatomical evaluation.

Impedance spectroscopy (IS) provides a rapid objective bedside assessment and has shown diagnostic performance for OASI detection when compared with EAUS in a prospective multicenter study.^9^ Whether IS provides additional clinically useful information when used alongside routine examination in the immediate postpartum setting has not been established. We therefore aimed to evaluate the added diagnostic value of immediate postpartum IS when used alongside routine clinical examination for detecting OASI, using subsequent EAUS as the anatomical reference assessment.

## Methods

### Study design

This prospective observational cohort study evaluated IS alongside routine postpartum clinical assessment following vaginal birth. The study was conducted at Holy Family Specialist Hospital, Warsaw, Poland, a high-volume obstetric center with approximately 3,700 deliveries annually, of which approximately 59% are vaginal. Women meeting the predefined eligibility criteria were prospectively enrolled between October 2025 and May 2026.

The study was approved by the Bioethics Committee at the District Medical Chamber in Warsaw (approval no. RNN/312/25/KE). All participants provided written informed consent before enrollment. The study was not registered. The study protocol is not publicly available.

### Study population

Women aged ≥18 years following vaginal birth were eligible if they had at least one predefined risk factor for OASI - primiparity, operative vaginal birth, neonatal birthweight ≥4000 g or gestational age ≥41 weeks - or if the postpartum clinical assessment raised uncertainty regarding possible sphincter injury.^4^

Exclusion criteria were inability to provide informed consent, clinical instability following delivery, or a contraindication to rectal assessment, including EAUS, as determined by the treating clinician.

Eligible women were approached consecutively during the study period when clinical circumstances permitted recruitment. Enrollment was limited to women who agreed to participate and provided written informed consent.

### Immediate postpartum clinical assessment

Following vaginal birth, participants underwent clinical assessment according to local departmental practice, including visual examination of the perineum and DRE to assess anal sphincter integrity. Where the initial examination raised uncertainty regarding possible OASI, the woman underwent further assessment by a senior obstetrician.

IS was performed during the immediate postpartum period and the result was available to the treating obstetrician as additional objective information. Clinical management was based on the available clinical findings and IS result. Where these findings were discordant, the final decision regarding the diagnosis of OASI and primary sphincter repair remained with the treating obstetrician.

### Impedance spectroscopy

IS was performed using the ONIRY® system (OASIS Diagnostics, Warsaw, Poland), comprising a dedicated single-use anal probe connected to an impedance spectrometer. The probe acquires frequency-dependent electrical impedance measurements from the perianal tissues, which are analyzed using a predefined machine-learning algorithm.

Measurements were performed by physicians and midwives trained in the use of the ONIRY system, according to the standard examination procedure. The predefined algorithm generated a binary PASS or REFER result, corresponding respectively to a negative or positive IS result for OASI.

The technical principles of IS, the measurement procedure and development of the machine-learning classification model have been described previously.^9–11^

### Reference assessment

Three-dimensional endoanal ultrasound (3D EAUS) was planned as the reference assessment for all enrolled women and was performed subsequently in a dedicated perineal clinic within the same hospital. Of the 151 women enrolled, 70 returned for the planned EAUS follow-up and were included in the paired diagnostic analysis; the remaining 81 did not attend the scheduled EAUS examination. Examinations were performed by a single expert ultrasonographer with more than 25 years of experience in anal sphincter imaging, using a BK Medical ultrasound system (BK Medical, Denmark) equipped with a dedicated 16 MHz and 360° rotational endoanal transducer for three-dimensional image acquisition.

The EAS and IAS were assessed separately for anatomical integrity and evidence of sphincter injury, including sphincter defects and post-injury or post-repair appearances.^3,6^ In women who had undergone primary sphincter repair, EAUS assessment also included postoperative sphincter anatomy, the presence and extent of any sphincter defect, and sonographic features of recent repair, including visible suture material and scar formation. These features allowed recent repaired OASI related to the index delivery to be distinguished from intact sphincter anatomy, including where no sphincter defect was present.

EAUS findings were classified as intact sphincter anatomy, unrepaired OASI or repaired OASI, with EAS and IAS involvement assessed separately. For the diagnostic analyses, both unrepaired and repaired OASI were classified as OASI-positive, while intact sphincter anatomy without evidence of OASI related to the index delivery was classified as OASI-negative. The EAUS examiner was not blinded to the findings of the clinical assessment or IS.

### Outcome measures

The primary outcome was the proportion of women with EAUS-confirmed OASI not identified by clinical assessment who generated an IS REFER result.

Secondary outcomes included the diagnostic performance and agreement of clinical assessment and IS against EAUS, and the frequency and characteristics of clinically relevant discordant findings.

### Statistical analysis

The predefined OASI risk factors were primiparity, operative vaginal birth, neonatal birthweight ≥4000 g and gestational age ≥41 weeks, based on contemporary international guidance.^4^ The number of risk factors present in each participant was also determined.

Continuous variables were summarized as mean ± SD or median (IQR), as appropriate according to their distribution. Categorical variables were presented as frequencies and percentages. No formal sample-size calculation was performed. Study size was determined pragmatically by the prospectively enrolled cohort available for the present analysis.

Diagnostic analyses were restricted to the 70 women who completed the planned EAUS assessment and had complete clinical assessment, IS and EAUS data. Diagnostic performance of clinical assessment and IS was evaluated against the EAUS reference assessment using sensitivity, specificity, positive predictive value, negative predictive value and overall diagnostic accuracy, with 95% CIs calculated using the Wilson method. Agreement of clinical assessment and IS with the EAUS reference assessment was evaluated using Cohen’s κ coefficient. Diagnostic accuracy of clinical assessment and IS was compared using the exact McNemar test for paired data.

Discordant findings between the clinical assessment and EAUS were examined descriptively. IS results were evaluated particularly in women with EAUS-confirmed OASI not identified clinically and in women clinically diagnosed with OASI who underwent primary repair but had no sphincter defect on subsequent EAUS. The distribution of predefined OASI risk factors within these groups was also described.

Statistical analyses were performed using Python version 3.10.8. A two-sided P value <0.05 was considered statistically significant.

## Results

### Study population

During the study period, 1432 vaginal births occurred and 151 women were enrolled. All enrolled women underwent immediate postpartum clinical assessment and IS. Of the 151 enrolled women, 70 (46.4%) returned for the planned EAUS follow-up and had complete clinical assessment, IS and EAUS data; these women comprised the paired diagnostic cohort. The remaining 81 women (53.6%) did not attend the scheduled EAUS examination and were therefore not included in the paired diagnostic analysis (Figure 1).

**Figure 1.**
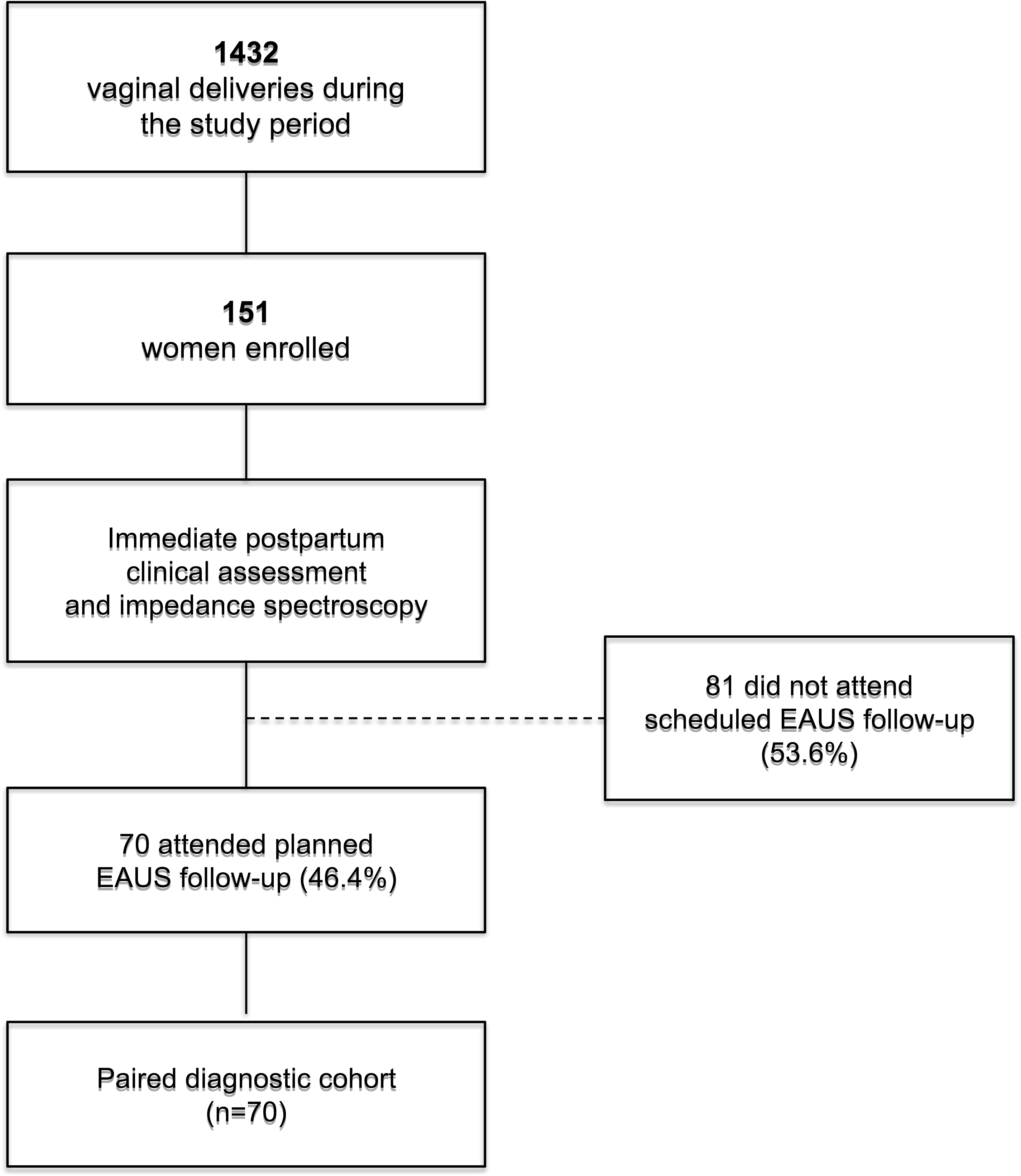
Study flow diagram. EAUS, endoanal ultrasound.

Among the 70 women included in the paired diagnostic cohort, 52 (74.3%) had at least one predefined OASI risk factor. Maternal, obstetric and delivery characteristics are presented in Table 1.

**Table 1.** Maternal, obstetric and delivery characteristics of the study cohort.

| Characteristic | Value |
| --- | --- |
| Maternal and obstetric characteristics |  |
| Maternal age, years, median (IQR) | 32.5 (29.0–35.0) |
| Gestational age at delivery, weeks, median (IQR) | 39 (39–40) |
| Neonatal birthweight, g, median (IQR) | 3580 (3276–3909) |
| Primiparous, n (%) | 42 (60.0) |
| Operative vaginal birth, n (%) | 11 (15.7) |
| └ Vacuum | 10 (14.3) |
| └ Forceps | 1 (1.4) |
| Neonatal birthweight $\geq 4000$ g, n (%) | 14 (20.0) |
| Gestational age $\geq 41$ weeks, n (%) | 8 (11.4) |
| Number of predefined OASI risk factors, n (%) |  |
| None | 18 (25.7) |
| One | 31 (44.3) |
| Two | 19 (27.1) |
| Three | 2 (2.9) |
| $\geq 1$ risk factor | 52 (74.3) |
| Timing and performance of diagnostic assessments |  |
| Time from delivery to IS, hours, median (range) | 1 (1–4) |
| IS performed within first hour postpartum, n (%) | 66 (94.3) |
| Technically valid IS recording, n (%) | 70 (100) |
| IS performed by midwife, n (%) | 49 (70.0) |
| IS performed by physician, n (%) | 21 (30.0) |
| Time from delivery to EAUS, days, median (IQR) | 28 (19.25–31.75) |
| Time from delivery to EAUS, days, range | 1–40 |

Footnote: Predefined OASI risk factors were primiparity, operative vaginal birth, neonatal birthweight
$\geq 4000$ g and gestational age $\geq 41$ weeks.
Abbreviations: EAUS, endoanal ultrasound; IQR, interquartile range; IS, impedance spectroscopy;
OASI, obstetric anal sphincter injury.

IS was performed within the first hour postpartum in 66/70 women (94.3%), and all recordings were technically valid. Most examinations were performed by midwives (49/70, 70.0%). EAUS was performed at a median of 28 days postpartum (IQR 19.25–31.75; range 1–40 days). No adverse events related to IS or EAUS were reported.

### Clinical and EAUS findings

Clinical assessment classified 34/70 women (48.6%) as having OASI and 36/70 (51.4%) as having no OASI. Among the 34 clinically identified injuries, 20 were classified as grade 3a, 10 as grade 3b, three as grade 3c and one as a fourth-degree injury. IS generated a REFER result in 28/70 women (40.0%) and a PASS result in 42/70 (60.0%).

EAUS-based reference assessment classified 29/70 women (41.4%) as OASI-positive and 41/70 (58.6%) as OASI-negative. Among the 29 women with confirmed OASI, 25 had injury involving the EAS alone and four had combined EAS and IAS injury. Twenty-two of the 29 OASI had been recognized clinically and repaired before EAUS, whereas seven had not been identified during the clinical assessment and had not undergone sphincter repair.

### Diagnostic performance

Compared with the EAUS-based reference assessment, clinical assessment correctly identified 22/29 women with OASI and correctly classified 29/41 women without OASI. Seven confirmed OASI were not identified clinically, while 12 women with a positive clinical assessment were classified as OASI-negative on the EAUS-based reference assessment. Clinical assessment had a sensitivity of 75.9% (95% CI 57.9–87.8), specificity of 70.7% (95% CI 55.5–82.4), PPV of 64.7% (95% CI 47.9–78.5), NPV of 80.6% (95% CI 65.0–90.2), and overall accuracy of 72.9% (95% CI 61.5–81.9).

IS correctly identified 25/29 women with confirmed OASI and correctly classified 38/41 women without OASI. Four women with confirmed OASI generated a PASS result and three OASI-negative women generated a REFER result. IS had a sensitivity of 86.2% (95% CI 69.4–94.5), specificity of 92.7% (95% CI 80.6–97.5), PPV of 89.3% (95% CI 72.8–96.3), NPV of 90.5% (95% CI 77.9–96.2), and overall accuracy of 90.0% (95% CI 80.8–95.1) (Table 2).

**Table 2.** Diagnostic performance of clinical assessment and IS against the EAUS-based reference assessment.

| Measure | Clinical assessment | IS |
| --- | --- | --- |
| True positive, n | 22 | 25 |
| False negative, n | 7 | 4 |
| True negative, n | 29 | 38 |
| False positive, n | 12 | 3 |
| Sensitivity, % (95% CI) | 75.9 (57.9–87.8) | 86.2 (69.4–94.5) |
| Specificity, % (95% CI) | 70.7 (55.5–82.4) | 92.7 (80.6–97.5) |
| PPV, % (95% CI) | 64.7 (47.9–78.5) | 89.3 (72.8–96.3) |
| NPV, % (95% CI) | 80.6 (65.0–90.2) | 90.5 (77.9–96.2) |
| Accuracy, % (95% CI) | 72.9 (61.5–81.9) | 90.0 (80.8–95.1) |

Abbreviations: CI, confidence interval; EAUS, endoanal ultrasound; IS, impedance spectroscopy;
NPV, negative predictive value; PPV, positive predictive value.

### Agreement with the EAUS-based reference assessment

Agreement with EAUS was higher for IS than for clinical assessment. IS and EAUS gave concordant classifications in 63/70 women (90.0%; Cohen’s κ=0.79), compared with 51/70 (72.9%; κ=0.45) for clinical assessment and EAUS.

In the paired comparison of diagnostic accuracy, IS was correct and clinical assessment incorrect in 17 women, whereas clinical assessment was correct and IS incorrect in five. This difference was statistically significant (exact McNemar P=0.017).

### Discordant diagnostic findings

Seven of the 29 women with EAUS-confirmed OASI had no OASI identified during clinical assessment. All seven generated an immediate postpartum IS REFER result. Three of these seven women had none of the four predefined OASI risk factors. Patterns of agreement and discordance between clinical assessment, IS and the EAUS-based reference assessment are summarized in Table 3. Overall, four women with EAUS-confirmed OASI generated an IS PASS result. All four had been identified as having OASI during clinical assessment and underwent primary sphincter repair.

**Table 3.** Patterns of agreement and discordance between clinical assessment, IS and the EAUS-based reference assessment.

| Clinical assessment | IS result | EAUS-based reference assessment | n |
| --- | --- | --- | --- |
| Positive | REFER | OASI-positive | 18 |
| Negative | PASS | OASI-negative | 28 |
| Negative | REFER | OASI-positive | 7 |
| Positive | PASS | OASI-positive | 4 |
| Positive | REFER | OASI-negative | 2 |
| Negative | REFER | OASI-negative | 1 |
| Positive | PASS | OASI-negative | 10 |
| Total |  |  | 70 |

Abbreviations: EAUS, endoanal ultrasound; IS, impedance spectroscopy; OASI, obstetric anal sphincter injury.

Twelve women had a positive clinical assessment for OASI but were classified as OASI-negative on the EAUS-based reference assessment. All 12 underwent primary sphincter repair before EAUS. IS generated a PASS result in 10/12 women (83.3%) and a REFER result in two (16.7%).

Three OASI-negative women generated an IS REFER result. Two had a positive clinical assessment for OASI and underwent primary sphincter repair, while one had no OASI identified clinically.

## Discussion

### Principal findings

In this prospective observational study, immediate postpartum IS provided additional objective information alongside clinical assessment for OASI. IS showed greater agreement with the EAUS-based reference assessment than clinical assessment (90.0% vs 72.9%) and higher diagnostic accuracy in the paired analysis.

The clinically most relevant findings were the patterns of discordance. Seven women with confirmed OASI were not identified through the clinical assessment pathway, despite senior reassessment when the initial examination was uncertain; all seven generated an immediate postpartum IS REFER result. Conversely, 12 women were clinically diagnosed with OASI and underwent primary sphincter repair but were subsequently classified as OASI-negative on the EAUS-based reference assessment; 10 had generated an IS PASS result. Importantly, this classification considered sonographic evidence of recent injury and repair and was not based solely on the absence of a residual sphincter defect.

These findings suggest that the potential contribution of IS may lie particularly where clinical assessment does not provide sufficient confidence to identify or exclude sphincter injury.

### Clinical interpretation and previous evidence

Accurate recognition of OASI immediately after birth can be difficult because clinical assessment requires both identification and interpretation of disrupted sphincter anatomy. Disagreement between clinical and ultrasound assessment has been reported across different levels of obstetric experience, and previous imaging studies have demonstrated both clinically unrecognized injury and discrepancies in injury classification, particularly involving the IAS.^1–3,6,12^

Previous diagnostic studies have shown that IS can distinguish injured from non-injured sphincter tissue when evaluated against EAUS.^9–11^ The present findings extend this evidence to the immediate postpartum clinical pathway. To our knowledge, this is the first prospective real-world evaluation focused specifically on IS in this setting, with almost all examinations performed within the first hour after birth, when decisions regarding recognition, primary repair and further assessment are still being made.

### Role of EAUS and place of IS in the clinical pathway

The potential clinical value of an additional objective test is likely to lie in the diagnostic grey zone rather than in clinically obvious OASI. When sphincter injury is clearly identified, established management should proceed without delay.⁴ A REFER result is not a diagnosis of OASI, and a PASS result should not override clear clinical evidence of injury. Rather, IS may provide additional objective information when clinical assessment does not provide sufficient confidence about sphincter integrity. The discordant findings support this complementary role. All four women with confirmed OASI who generated an IS PASS result had been identified clinically, whereas IS generated REFER in all seven women with OASI not identified clinically. Thus, no woman with EAUS-confirmed OASI in this cohort was negative on both clinical assessment and IS. A conceptual pathway illustrating this potential complementary role is shown in Figure 2.

**Figure 2.**
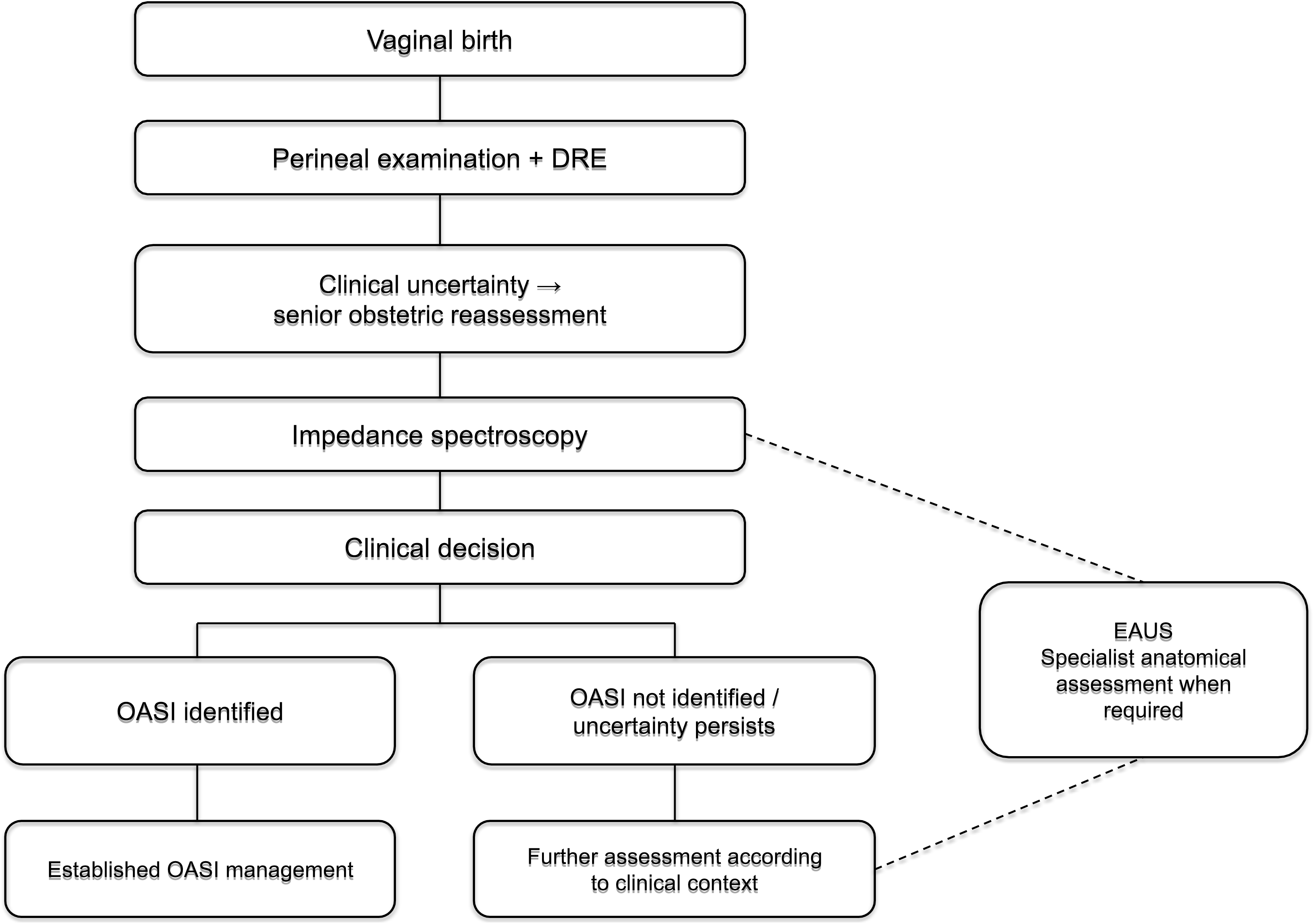
Conceptual postpartum clinical assessment pathway incorporating impedance spectroscopy. The pathway illustrates the potential role of IS suggested by the present findings and is not a validated management algorithm. DRE, digital rectal examination; EAUS, endoanal ultrasound; IS, impedance spectroscopy; OASI, obstetric anal sphincter injury.

EAUS and IS therefore have different roles in the assessment of suspected OASI. EAUS provides detailed anatomical assessment of the EAS and IAS when precise evaluation is required,^4,7,13^ whereas IS provides a rapid bedside PASS/REFER result without defining the anatomical extent of injury.

Immediate postpartum EAUS requires dedicated equipment and expertise and may be challenging to interpret in the acute setting.^8,14^ IS may therefore provide additional objective information at the bedside, helping clinicians identify women in whom further clinical assessment or EAUS-based anatomical evaluation is warranted.

IS was performed within the first hour after birth in 94.3% of women; all recordings were technically valid and 70.0% were performed by midwives. This supports feasibility within routine maternity care, although operator-specific performance was not evaluated. Multidisciplinary working and staff engagement are important to effective OASI care.^15^ Potential effects on use of specialist imaging resources, efficiency and costs require prospective evaluation.

### Strengths and limitations

Strengths include prospective evaluation within routine immediate postpartum care and paired clinical assessment, IS and expert 3D EAUS, allowing assessment of both diagnostic performance and clinically relevant discordance.

Several limitations should be considered. This was a single-center study, and only 70 of 151 enrolled women completed EAUS, creating potential selection bias. Participants were selected because of predefined OASI risk factors or clinical uncertainty, resulting in a high OASI prevalence; predictive values should therefore not be extrapolated to an unselected postpartum population. The final cohort was also relatively small, limiting precision and subgroup analyses.

EAUS was performed subsequently rather than contemporaneously with clinical assessment and IS. Although expert 3D EAUS allowed assessment of sphincter integrity and sonographic features of recent injury and repair, the interval between delivery and imaging should be considered when interpreting discordant findings. In addition, IS results were available to treating clinicians and could influence decisions regarding reassessment and primary repair. This reflects real-world use but prevents determination of how management or subsequent EAUS findings would have differed without IS. The EAUS examiner was not blinded to earlier findings, introducing potential interpretation bias.

Finally, the study evaluated diagnostic findings rather than patient outcomes and cannot establish whether incorporating IS reduces missed OASI or inappropriate intervention, or improves follow-up and functional outcomes.

### Clinical implications and future research

Future studies should evaluate the complete PASS/REFER pathway rather than diagnostic accuracy alone, including detection of previously unrecognized OASI before discharge, referral for confirmatory assessment, changes in management and missed injury following a PASS result. Broader maternity populations and different professional groups should be included to establish generalizability.

The present findings also question whether future pathways should rely solely on risk-based selection: three of the seven women with clinically unrecognized OASI who generated REFER had none of the predefined risk factors. Broader postpartum assessment therefore warrants evaluation alongside targeted strategies.

Ultimately, diagnostic pathways should be judged by outcomes that matter to women. Given the association between OASI and long-term bowel dysfunction,^16^ future studies should determine whether improved recognition leads to appropriate treatment and follow-up, better bowel function and quality of life, while also evaluating feasibility, resource use and health-economic outcomes.

## Conclusion

Immediate postpartum impedance spectroscopy provided additional objective information alongside clinical assessment for OASI, particularly when clinical findings did not reliably identify or exclude sphincter injury. These findings support prospective evaluation of IS within complete postpartum diagnostic pathways to determine whether it reduces diagnostic errors and improves clinically meaningful outcomes.

## Use of generative artificial intelligence

During manuscript preparation, the authors used ChatGPT (GPT-5.6 Sol; OpenAI, L.L.C., San Francisco, CA, USA) for language refinement and light editorial assistance. The authors critically reviewed and revised all suggested changes and take full responsibility for the scientific content and final wording of the manuscript.

## Data Availability Statement

The data that support the findings of this study are available from the corresponding author upon reasonable request, subject to applicable ethical and data-protection requirements.

## STROBE Statement - Checklist for Cohort Studies

Improving Immediate Postpartum Detection of Obstetric Anal Sphincter Injury: Added Diagnostic Value of Impedance Spectroscopy Following Clinical Assessment Uchman-Musielak et al. | Ultrasound in Obstetrics & Gynecology (UOG)

Completed for UOG submission. Page numbers refer to the current submission manuscript.

| Section | Item | STROBE recommendation | Page No. | Location / manuscript note |
| --- | --- | --- | --- | --- |
| Title and abstract | 1a | Indicate the study design with a commonly used term in the title or abstract. | 2 | Abstract: prospective observational cohort study. |
| Title and abstract | 1b | Provide an informative and balanced abstract summary of what was done and found. | 2 | Structured Abstract: Objective, Methods, Results, Conclusions. |
| Introduction | 2 | Explain the scientific background and rationale for the investigation. | 3 | Introduction. |
| Introduction | 3 | State specific objectives, including any prespecified hypotheses. | 3 | Final paragraph of Introduction. |
| Methods | 4 | Present key elements of study design early in the paper. | 4 | Methods - Study design. |
| Methods | 5 | Describe setting, locations and relevant dates, including recruitment and follow-up. | 4-7 | Study design; Reference assessment; follow-up timing in Results. |
| Methods | 6a | Give eligibility criteria and sources/methods of participant selection; describe follow-up. | 4-5 | Study population; consecutive approach; planned EAUS follow-up. |
| Methods | 6b | For matched studies, give matching criteria and numbers. | N/A | Not a matched study. |
| Methods | 7 | Clearly define outcomes, exposures/predictors/confounders/effect modifiers and diagnostic criteria, if applicable. | 4-6 | Study population; Reference assessment; Outcome measures. |
| Methods | 8 | For each variable of interest, give data sources and measurement methods. | 4-6 | Clinical assessment, IS, EAUS and statistical analysis. |
| Methods | 9 | Describe efforts to address potential sources of bias. | 5-6, 11 | Reference-standard assessment/blinding is reported; limitations discuss selection and reference-assessment bias. |
| Methods | 10 | Explain how the study size was arrived at. | 6 | Statistical analysis.<br>No formal sample-size calculation was performed; study size was determined pragmatically by the prospectively enrolled cohort available for the present analysis. |
| Methods | 11 | Explain how quantitative variables were handled; describe groupings and rationale where applicable. | 4,6 | Predefined risk-factor thresholds and statistical analysis. |
| Methods | 12a | Describe all statistical methods, including methods used to control for confounding. | 6 | Statistical analysis; no adjusted/confounding model was performed. |
| Methods | 12b | Describe methods used to examine subgroups and interactions. | 6, 8 | Discordant groups and risk-factor distributions were examined descriptively; no interaction analysis. |
| Methods | 12c | Explain how missing data were addressed. | 5-6 | Paired diagnostic analyses restricted to 70 women with complete clinical assessment, IS and EAUS data. |
| Methods | 12d | If applicable, explain how loss to follow-up was addressed. | 5, 7, 11 | 81/151 did not attend EAUS follow-up; analysis restricted to completers; selection-bias implication discussed. |
| Methods | 12e | Describe any sensitivity analyses. | N/A | No sensitivity analyses were performed. |
| Results | 13a | Report numbers of individuals at each stage of the study. | 7; Fig. 1 | 1432 vaginal births; 151 enrolled; 70 attended EAUS; 81 did not attend; n=70 paired cohort. |
| Results | 13b | Give reasons for non-participation at each stage. | 7; Fig. 1 | 81 women did not attend the scheduled EAUS follow-up. Individual reasons were not systematically collected. |
| Results | 13c | Consider use of a flow diagram. | Fig. 1 | Study flow diagram. |
| Results | 14a | Give characteristics of study participants and relevant clinical/obstetric information. | 7; Table 1 (p. 15) | Study population and Table 1. |
| Results | 14b | Indicate number of participants with missing data for each variable of interest. | 6-7 | Paired analysis required complete clinical assessment, IS and EAUS data (n=70). |
| Results | 14c | Summarise follow-up time. | 7 | EAUS median 28 days postpartum (IQR 19.25-31.75; range 1-40). |
| Results | 15 | Report numbers of outcome events or summary measures over time. | 7-8 | Clinical/EAUS findings, diagnostic performance and discordant findings. |
| Results | 16a | Give estimates and their precision (e.g. 95% CI); adjusted estimates if applicable. | 8; Table 2 (p. 17) | Sensitivity, specificity, PPV, NPV and accuracy with 95% CIs; no adjusted estimates applicable. |
| Results | 16b | Report category boundaries when continuous variables were categorized. | 4,6 | Risk-factor thresholds are explicitly defined. |
| Results | 16c | If relevant, translate relative-risk estimates into absolute risk. | N/A | No relative-risk analysis. |
| Results | 17 | Report other analyses performed, including subgroup/interaction/sensitivity analyses. | 8; Table 3 (p. 18) | Agreement, paired McNemar comparison and descriptive discordance analyses. |
| Discussion | 18 | Summarise key results with reference to study objectives. | 9 | Principal findings. |
| Discussion | 19 | Discuss limitations, including potential bias/imprecision and direction/magnitude where possible. | 11 | Strengths and limitations. |
| Discussion | 20 | Give a cautious overall interpretation considering objectives, limitations and other evidence. | 9-12 | Clinical interpretation; role of EAUS/IS; limitations; implications. |
| Discussion | 21 | Discuss generalisability (external validity). | 11-12 | Selected/high-prevalence cohort and need for broader multicentre validation. |
| Other information | 22 | Give source of funding and role of funders. | 12 | Funding statement: no external funding; hospital costs; investigator-owned/provided ONIRY system and probes. |

